# Growth rate and acceleration analysis of the COVID-19 pandemic reveals the effect of public health measures in real time

**DOI:** 10.1101/2020.03.30.20047688

**Authors:** Yuri Tani Utsunomiya, Adam Taiti Harth Utsunomiya, Rafaela Beatriz Pintor Torrecilha, Silvana de Cássia Paula, Marco Milanesi, José Fernando Garcia

**Affiliations:** Department of Support, Production and Animal Health, School of Veterinary Medicine of Araçatuba, São Paulo State University (Unesp), 16050-680 R. Clovis Pestana 793 - Dona Amelia, Araçatuba/SP Brazil; International Atomic Energy Agency (IAEA) Collaborating Centre on Animal Genomics and Bioinformatics, 16050-680 R. Clovis Pestana 793 - Dona Amelia, Araçatuba/SP, Brazil; Department of Preventive Veterinary Medicine and Animal Reproduction, School of Agricultural and Veterinarian Sciences, São Paulo State University (Unesp), 14884-900 Via de Acesso Prof. Paulo Donato Castellane s/n - Jaboticabal/SP, Brazil

**Keywords:** Coronavirus, Severe Acute Respiratory Syndrome, Growth Curve Analysis, Mathematical Modeling, Moving Regression, Hidden Markov Model

## Abstract

**Background:** Ending the COVID-19 pandemic is arguably one of the most prominent challenges in recent human history. Following closely the growth dynamics of the disease is one of the pillars towards achieving that goal.

**Objective:** We aimed at developing a simple framework to facilitate the analysis of the growth rate (cases/day) and growth acceleration (cases/day^2^) of COVID-19 cases in real-time.

**Methods:** The framework was built using the Moving Regression (MR) technique and a Hidden Markov Model (HMM). The dynamics of the pandemic was initially modeled via combinations of four different growth stages: lagging (beginning of the outbreak), exponential (rapid growth), deceleration (growth decay) and stationary (near zero growth). A fifth growth behavior, namely linear growth (constant growth above zero), was further introduced to add more flexibility to the framework. An *R Shiny* application was developed, which can be accessed at http://www.theguarani.com.br/covid-19 or downloaded from https://github.com/adamtaiti/SARS-CoV-2. The framework was applied to data from the European Center for Disease Prevention and Control (ECDC), which comprised 853,200 cases reported worldwide as of April 2^nd^ 2020.

**Results:** We found that the impact of public health measures on the prevalence of COVID-19 could be perceived in seemingly real-time by monitoring growth acceleration curves. Restriction to human mobility produced detectable decline in growth acceleration within few days, deceleration within ∼2 weeks and near-stationary growth within ∼6 weeks. Countries exhibiting different permutations of the five growth stages indicated that the evolution of COVID-19 prevalence is more complex and dynamic than previously appreciated.

**Conclusions:** These results corroborate that mass social isolation is a highly effective measure against the dissemination of SARS-CoV-2, as previously suggested. Apart from the analysis of prevalence partitioned by country, the proposed framework is easily applicable to city, state, region and arbitrary territory data, serving as an asset to monitor the local behavior of COVID-19 cases.

## INTRODUCTION

The World Health Organization (WHO) officially declared Coronavirus Disease (COVID-19) a global pandemic on March 11^th^ 2020 (1). The disease is caused by the novel Severe Acute Respiratory Syndrome Coronavirus 2 (SARS-CoV-2) (2,3), which seems to have first emerged in Wuhan, China on December 12^th^ 2019 (4,5). Worldwide dissemination has been extremely rapid, and by the time this study was completed (April 2^nd^ 2020) a total of 928,437 cases and 46,891 deaths had been reported across 204 countries and territories according to data from the European Center for Disease Prevention and Control (ECDC) (6). Approximately 86% of all cases are estimated to have been undocumented prior to the cordon sanitaire in China (7), which suggests that the disease might be also substantially under-reported in other countries. Nevertheless, partial COVID-19 prevalence data are still an invaluable resource to help monitoring and controlling the disease. In particular, extracting daily estimates of growth rate (cases/day) and acceleration (cases/day^2^) in disease dissemination from real-time case reports can be decisive for an effective and promptly action to restrain further contagion. Here we report the development of a simple framework dedicated to the real-time analysis of COVID-19 prevalence. This framework was built using a combination of Moving Regression (MR) and Hidden Markov Model (HMM), and was deployed as a *Shiny* (8) application in *R* (9). Here we show the utility of that framework in the analysis of publicly available COVID-19 case reports that are updated daily by the ECDC.

## RESULTS AND DISCUSSION

For simplicity, assume that the cumulative number of COVID-19 cases over time (i.e., the growth curve) in a specific country or territory follows an unknown sigmoidal function (**Figure 1a**). Such assumption is common in the analysis of growth data and has been applied to a wide range of problems, from tumor (10) to bacterial (11) growth. Although empirical data from China (**Figure 1b**) and South Korea (**Figure 1c**) seemed to support it well, that assumption will be substantially relaxed later in our framework to accommodate complex dynamics in the evolution of COVID-19 prevalence. We define growth rate and growth acceleration as the first and second order derivatives, respectively, of the prevalence of COVID-19 in respect to time. In our framework, we selected MR to approximate these derivatives over competing models that are frequently used to describe the behavior of sigmoidal growth curves, such as the Gompertz model (12,13), because: (i) it is dependent on a single free parameter, the “smooth factor”, which represents the number of neighboring days used in local regression; (ii) growth rate and acceleration estimates are approximated by ordinary least squares equations, which are computationally inexpensive; (iii) we performed extensive simulations of growth curves and found that it produces reasonably accurate estimates of growth rate (median *R*^2^ = 0.99 with smooth factor of 3) and acceleration (median *R*^2^ = 0.92 with smooth factor of 3) (**Figure 2**); (iv) it is very robust to departures from sigmoidal curves; and (v) it does not rely on observations of the whole curve to produce instantaneous growth rate and acceleration estimates, and thus can produce such estimates in near real time. Argument (v) is especially relevant to the analysis of COVID-19 data since the pandemic is ongoing and each country will be at a different stage of the growth curve as time passes. A clear disadvantage of MR is that it may over-fit the growth curve to the data, especially if the selected smooth factor is small (say < 3), in which case accurate prediction of new cases of COVID-19 is limited to very few days in the future. Still, even single-day predictions can be of great use during a pandemic if reasonably accurate. In the ECDC data set, a forward validation showed that single-day predictions were sufficiently accurate (*R*^2^ ∼ 0.95) (**Figure 3**).

**FIGURE 1.**
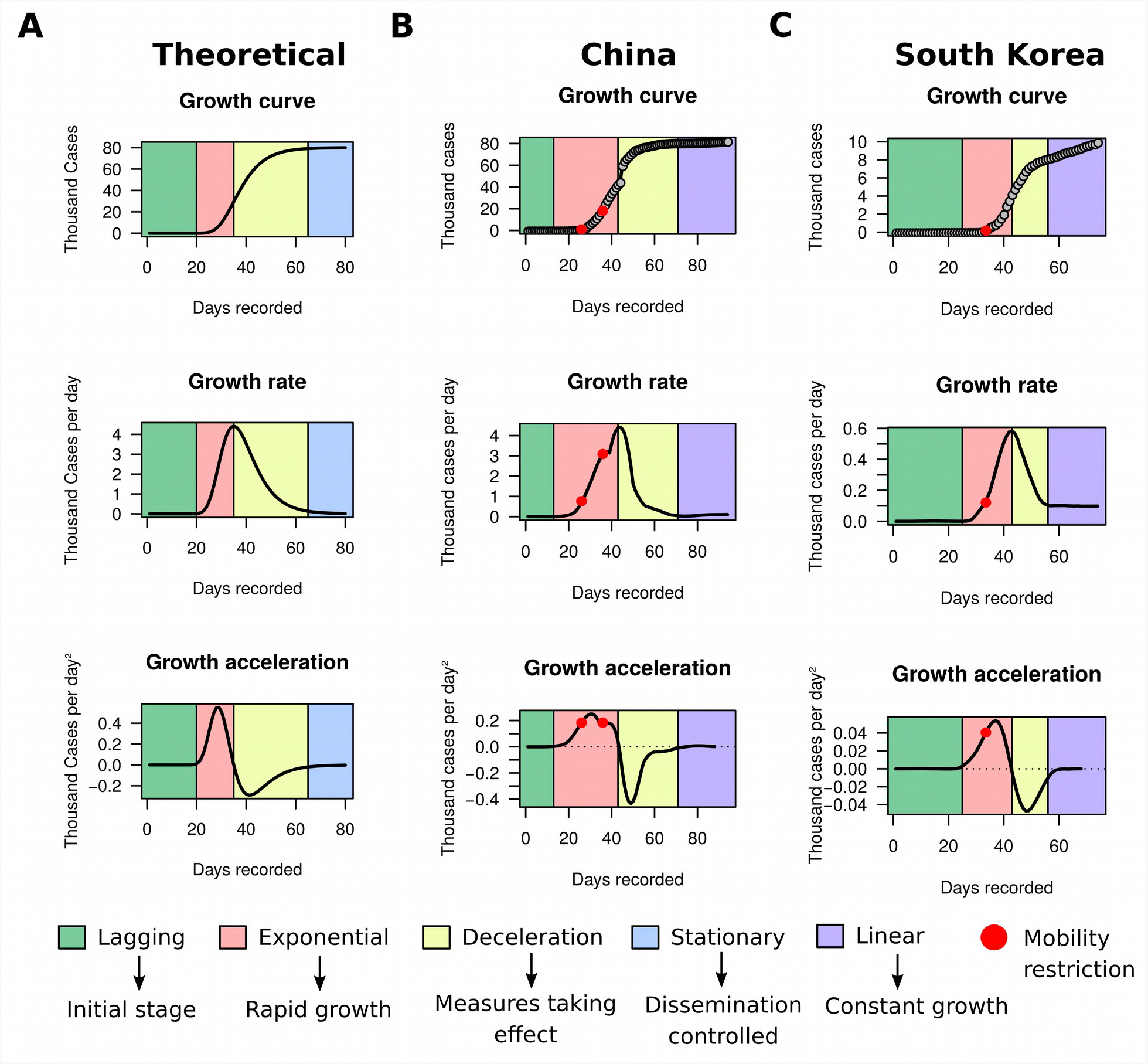
Growth rate and acceleration in China and South Korea. (**A**) Theoretical model exemplified by simulated data using a four-parameters Gompertz model with an asymptote at 80,000, growth coefficient of 0.15, inflection time at 35, and time ranging from 1 to 80. (**B**) Fitted curves for China between December 31^st^ 2019 and April 2^nd^ 2020. The first red dot marks the midpoint between January 23^rd^ and 24^th^ 2020, when a strict cordon sanitaire was imposed to Wuhan, Shanghai, Jiangsu and Hainan. The second red dot pinpoints February 4^th^ 2020, when the cordon was extended to a larger portion of the eastern part of China. (**c**) Fitted curves for South Korea between January 20^th^ and April 2^nd^ 2020. The red dot is placed between February 20^th^ and 21^st^ 2020, when a collection of restrictions to human mobility was imposed, including lockdown of Daegu city, suspension of flights, cancellation of mass gatherings and lockdown of all South Korean military bases. The apparent stationary stage of both countries is in reality classified as linear growth by our framework, since the approximately constant growth rate in this period is larger than the maximum growth rate observed during the lagging stage.

**FIGURE 2.**
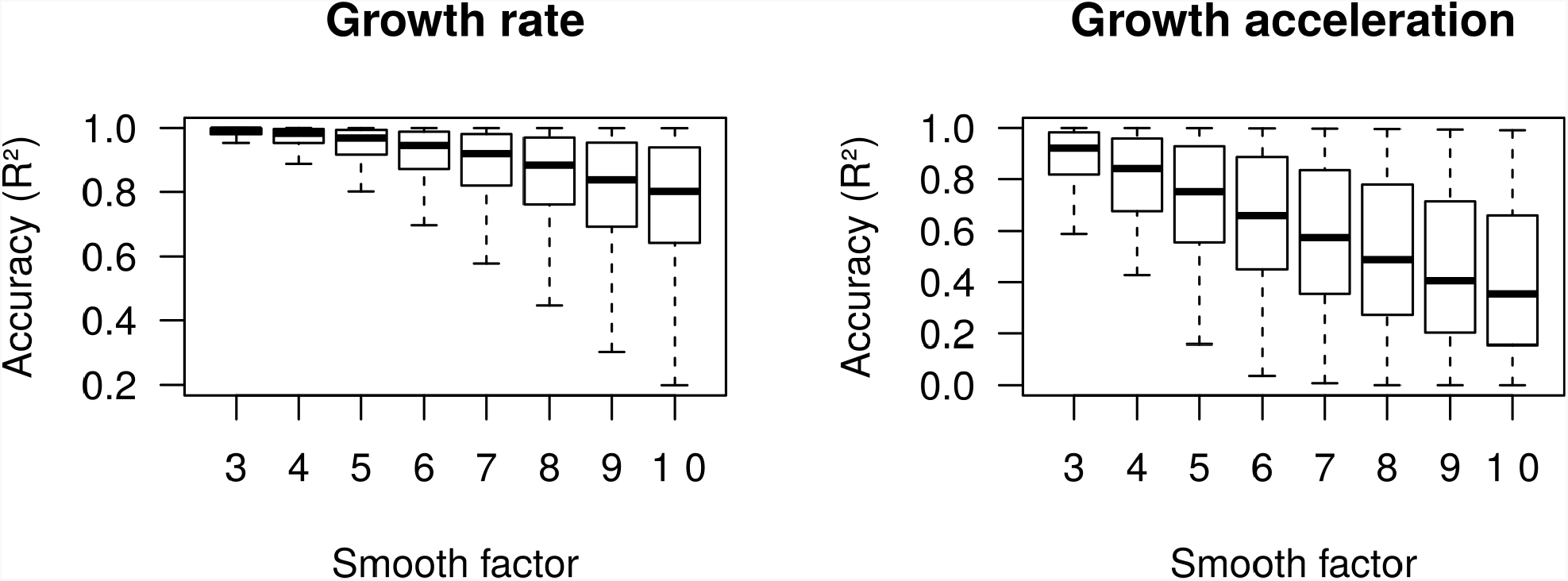
Accuracy (*R*^2^) of moving regression estimates of growth rate and growth acceleration from 50,000 simulated Gompertz growth curves.

**FIGURE 3.**
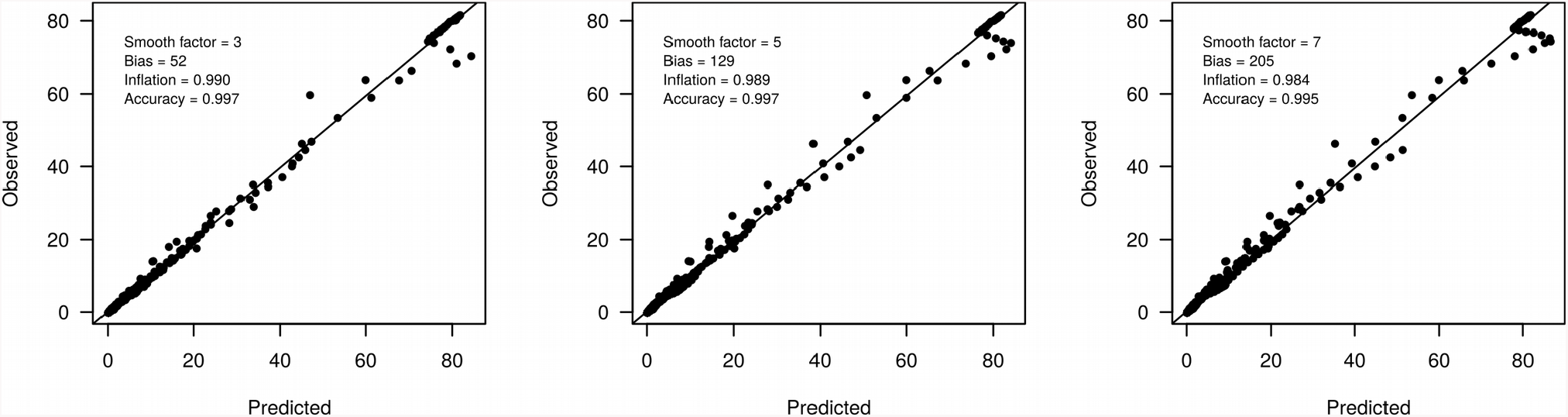
Accuracy (*R*^2^) of moving regression predictions of next-day COVID-19 prevalence.

Sigmoidal growth curves can be partitioned into four easily distinguishable stages (**Figure 1a**): (a) the lagging stage, which corresponds to the beginning of the outbreak or disease importation, where the number of cases are low and increase only marginally every day; (b) the exponential stage, when growth starts accelerating and the number of new cases increase rapidly day-by-day; (c) the deceleration stage, where the number of new cases reduces daily and tends to asymptote; and (d) the stationary stage, characterized by stagnation of the prevalence with sporadic new cases occurring each day. The growth rate graph is approximately bell-shaped, with its peak corresponding to the inflection of the exponential stage. This inflection point signals the beginning of a decline in the growth rate. The growth acceleration graph usually consists of a combination of two bell-shaped curves: the first one with a peak and the second with a valley. The peak indicates the point where acceleration starts descending towards zero. The moment when acceleration is exactly zero coincides with the inflection of the exponential stage, which marks the beginning of growth deceleration (i.e., negative acceleration). The latter corresponds to the entire concave section of the curve, but the very bottom of the valley indicates that the prevalence is moving towards stagnation.

In spite of sigmoidal curves following the four above described stages sequentially, we anticipated that the growth of COVID-19 cases may not necessarily obey this sequence in practice, since the dynamics of the disease is likely complex and highly responsive to the implementation or relaxation of public health measures. This implies that a country that has already reached a stationary stage could resume exponential growth, for example by seeding a new outbreak via importation. Likewise, decelerating countries could as well regain acceleration by relaxing prevention measures. Furthermore, some countries may face multiple cycles of acceleration and deceleration prior to reaching a stationary growth. These scenarios could produce more complex growth curves that deviate from the sigmoidal shape by mounting different arrangements of exponential, deceleration and stationary stages. Of note, MR has sufficient flexibility to model these complex scenarios and can easily accommodate curves exhibiting arbitrary permutations of these four stages. In addition, the near-zero acceleration that is intimately related to the stationary stage in sigmoidal curves could also arise from a non-zero constant growth rate in practice. In such cases, the growth curve would exhibit a linear pattern, which can be interpreted as a fifth growth stage that is not observed in classic sigmoidal functions. Such linear pattern may appear if the deceleration stage does not form an enough deep valley prior to acceleration rising up again towards zero. Again, MR is capable of modeling these anomalous behaviors. In this study we sought to ascertain whether these five stages of growth curves could have direct implications in understanding the dynamics of COVID-19 prevalence both globally and locally. We further developed a Hidden Markov Model (HMM) to automate the detection of transitions between stages in the growth curve using acceleration and growth rate data obtained with MR as input (see **Material and Methods**).

Using MR and HMM on ECDC data frozen on April 2^nd^ 2020, we evaluated the utility of the framework in identifying countries reaching deceleration or stationary growth. We also looked for countries presenting complex arrangements of the five growth stages. The countries found to have reached a near-stationary stage at some point were China and South Korea (**Figure 1b-c**). By projecting official government announcements against the fitted curves of these countries, we observed that decline in growth acceleration occurred shortly after the implementation of measures that drastically reduced human movement. Deceleration of growth was achieved within 2 weeks and the prevalence plateaued within 6 weeks. These results indicated that: (i) the effect of public health measures on SARS-CoV-2 prevention can be detected in seemingly real time by monitoring the behavior of acceleration curves; and (ii) restriction to human mobility is very effective in controlling the spread of the disease, but takes several weeks to produce a stationary growth. These findings are in line with a recent study showing that human mobility explained early growth and decline of new cases of COVID-19 in China (14). As discussed before, one should not immediately assume that a country in stationary growth will remain in that stage, since acceleration could take off again if new cases are imported or preventive measures are relaxed. In fact, our HMM classifier categorized the apparent stationary growth stage of China and South Korea as a linear growth. Indeed, both countries have not reached a perfect asymptote and their COVID-19 prevalences are instead growing in a linear pattern.

To illustrate the utility of the framework in detecting acceleration decay, we decided to look more closely to data from three countries: United States of America (USA), Brazil and Italy (**Figure 4**). The latter has been severely impacted with the disease, and by the time we completed our study the country had recorded 110,574 cases and 13,157 deaths. On March 10^th^ 2020 Italy implemented a strict quarantine, and five days later the country reached its maximum acceleration and started to move towards an inflection of the exponential growth. On March 25^th^ Italy further implemented a complete shut down of its borders, and our analysis showed that the country started to decelerate on March 26^th^. Brazil and USA both showed brief declines in acceleration between March 20^th^ and 25^th^, but were still found in exponential growth as of April 2^nd^. In fact, Brazil resumed acceleration incline on March 26^th^. Still, since acceleration response to effective measures seems rapid, both Brazil and USA could begin a deceleration process within few weeks if effective measures are implemented are rigorously followed. To date, Brazil has accumulated 6,836 cases and 241 deaths, whereas USA has recorded 216,721 cases and 5,138 deaths.

**FIGURE 4.**
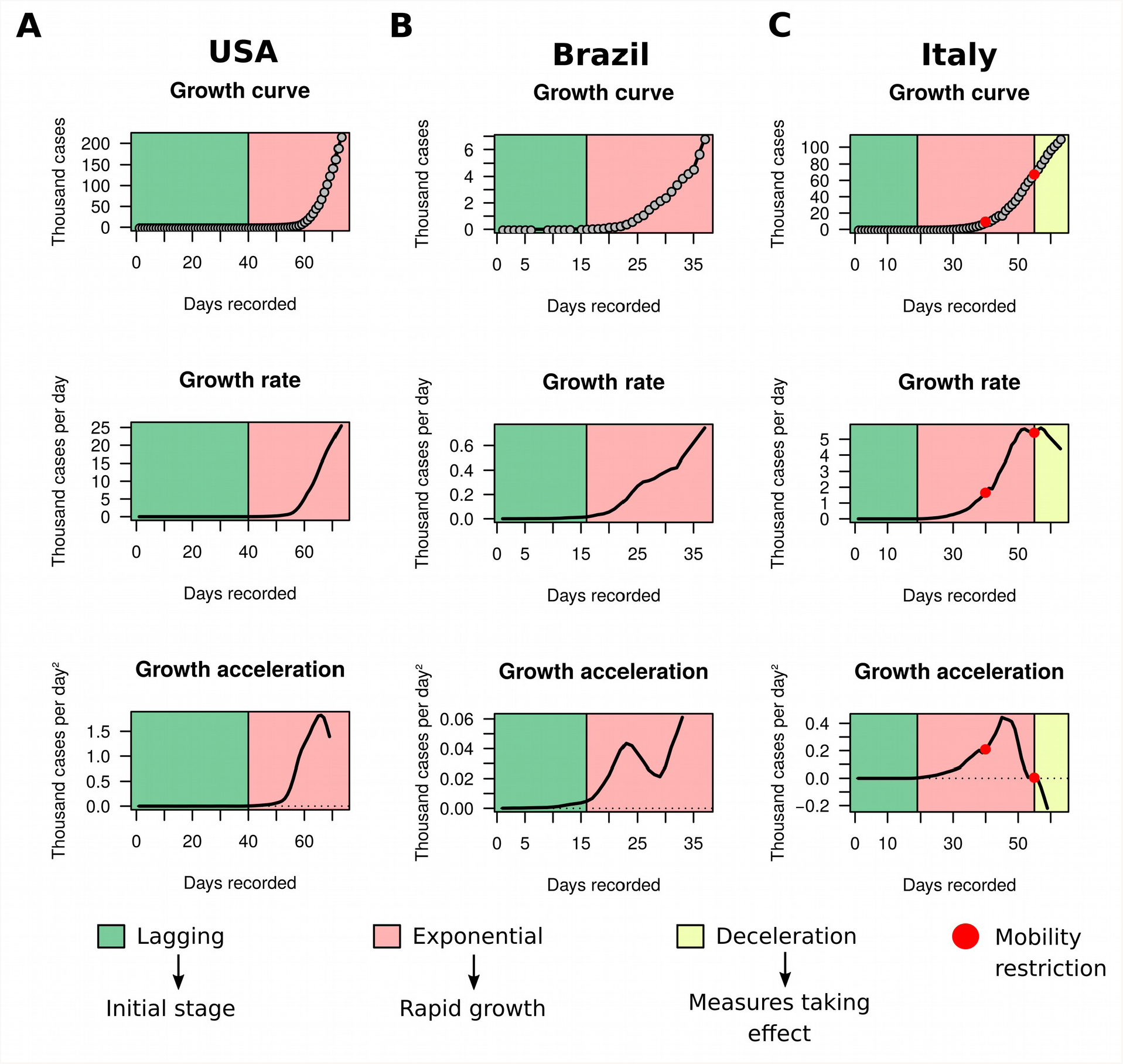
Growth rate and acceleration in the United States of America (USA), Brazil and Italy. By the time the study was completed, the governments of the United States of America (**a**) and Brazil (**b**) had not announced severe strict measures to restrain human movement, such as a lockdown. As a measure to reduce movement, these two countries had primarily focused on preventing mass gatherings, besides of closing schools, nurseries, universities and other places that facilitate agglomeration, such as shopping malls. This is in contrast with Italy (**c**), which imposed strict quarantine on March 10^th^ 2020 (first red dot) and closure of borders on March 25^th^ 2020 (second red dot). The country is currently in growth deceleration.

**FIGURE 5.**
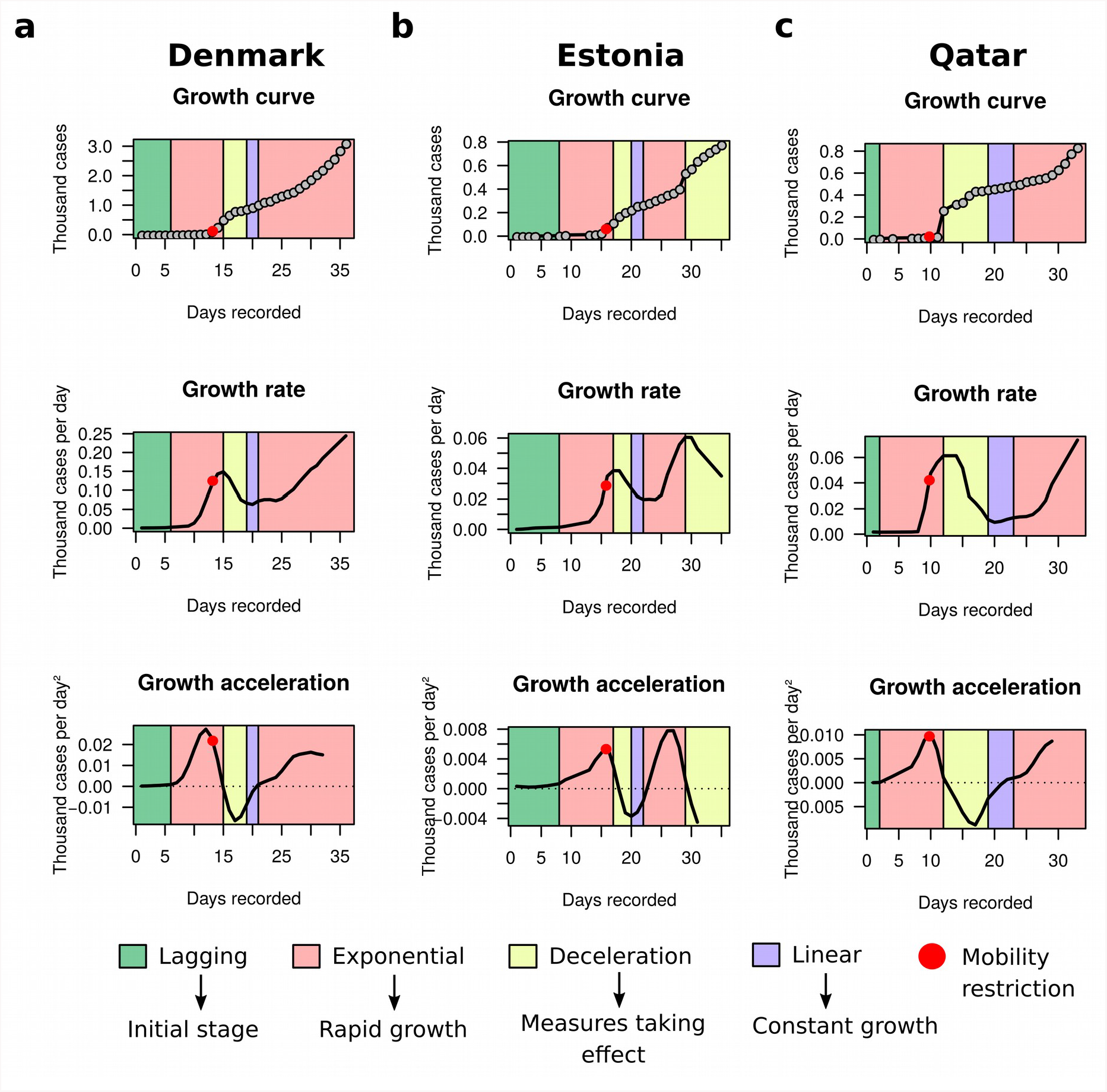
Growth rate and acceleration in Denmark, Estonia and Qatar. These three countries exhibited complex arrangements of growth stages. (**a**) On March 11^th^ 2020 (red dot), Denmark became the second European country to establish a lockdown. The country started decelerating new cases of COVID-19 two days after. (**b**) On March 13^th^ 2020 (red dot) a state of emergency was declared by the Estonian government, which imposed significant restrictions to travel and mass gatherings. Decline of growth rate was observed one day after, and deceleration started within 4 days. (**c**) Our analysis indicated that Qatar reached its peaking acceleration on March 9^th^ 2020 (red dot), the same day when Qatari officials had announced the closure of schools, nurseries, and universities, in addition to strong restrictions to traveling. One day after, the country registered a sudden spike in the number of cases (arrow). The days that followed were marked by a clear decline in acceleration and eventually deceleration of growth, which coincided with a succession of measures by the Qatari government that eventually led to significant restrictions to human movement. Although stationary growth was expected after deceleration, these three countries passed through a short linear phase and then regained acceleration, which initiated a new exponential phase. Estonia has further initiated a second deceleration stage on March 27^th^ 2020.

To exemplify the ability of our framework in detecting complex arrangements of growth stages that substantially deviate from the sigmoidal model, we selected Denmark, Estonia and Qatar (**Figure 4**). All three countries experienced deceleration phases that did not culminate in stationary growth. Instead, a brief linear growth was formed after deceleration, which was followed by a new exponential growth stage. Estonia has further entered a new deceleration phase on March 27^th^. These observations indicate that the growth dynamics of COVID-19 cases is more complex than previously appreciated. Therefore, analyzing the raw growth curve alone, dissociated from its derivatives, is very limiting for inference and may hamper the understanding of the pandemic evolution. In part, the lack of combined analysis of growth rate and acceleration in this pandemic is to be blamed on scarce availability of tailor made, user-friendly software. To aid to the analysis of growth rate and acceleration of COVID-19 cases, we built a web application using *R* (9) and *Shiny* (8). This application automatically loads the latest ECDC case reports and applies MR to extract growth rate and acceleration from real-time data. The app also performs automated classification of growth stages with HMM (albeit free parameters should be manually tuned for improved results). Users are not limited to case reports from ECDC, since the app allows for the upload of custom data (e.g., city, region, province or state), which can be used to monitor the growth behavior of COVID-19 locally. Upon closing of the COVID-19 pandemic, this tool could be further used in the analysis of future outbreaks and epidemics, or even of historical disease data. A limiting factor however is that the proposed framework relies on updated case reports, such that sub-notification, delayed communication and the elapsed time between sample collection, diagnostic results and reporting may impact the real-time inference of growth dynamics in disease transmission and consequently jeopardize the timely detection of transitions in the growth curve. In spite of that limitation, the presented tool remains highly useful to monitor the growth behavior of epidemics.

## CONCLUSIONS

We deployed a simple framework for the real-time analysis of COVID-19 prevalence. We were able to demonstrate that the real-time decomposition of growth curves of COVID-19 cases into growth rate and acceleration can be a powerful tool to monitor the impact of public health measures on the spread of the disease. We also showed that restrictions to human mobility can significantly decelerate the incidence of new cases within weeks. Furthermore, we found that the prevalence of the disease is more complex and dynamic than previously appreciated. This observation will have important implications to assumptions adopted in mathematical models to predict the evolution of the pandemic.

## MATERIAL AND METHODS

### Moving regression (MR) model

The MR technique adopted here aimed at fitting a smooth growth curve to the COVID-19 prevalence data, such that the resulting curve could describe the cumulative number of cases as a function of time. For *n* recorded days in a given country or territory, let **x** be a *n*-dimensional column vector of days since the first case report and **y** the reciprocal column vector with elements corresponding to the cumulative number of cases. Relative to day *d*, we define **y**_d_ and **x**_d_ as *k*-sized subset vectors of **y** and **x**, respectively, where *k* = 1 + 2*s* and *s* is a free parameter representing the number of offset days before and after day *d*. Hereafter, we refer to *s* as the “smooth factor”, since it controls the compromise between over-smoothing (large *s*) and over-fitting (small *s*) the curve to the data. Finally, we define **X**_d_ = [**1**_k_ **x**_d_], where **1**_k_ is a *k*-dimensional column vector with all elements equal to one. The local growth rate was estimated by ordinary least squares regression:

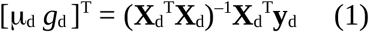

where μ_d_ is an intercept and *g*_d_ is the estimated growth rate (cases/day) at day *d*. In practice, *g*_d_ corresponds to an estimate of the instantaneous rate of change in the number of cases at day *d*, which in turn is an approximation to the first order derivative of the unknown growth function evaluated at time *d*. The smoothed growth curve was obtained by calculating fitted values as:

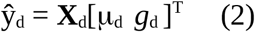

After fitting equation (1) to all *n* records, we define **g** is a vector of size *n* containing all estimated local growth rates and **g**_d_ as a *k*-sized subset vector of **g**. The local growth acceleration at day *d* was then obtained by adapting equation (1):

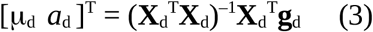

where *a*_d_ is the estimated growth acceleration (cases/day^2^) at day *d*. Now *a*_d_ is an estimate of the instantaneous rate of change of the growth rate at day *d*, which consequently approximates the second order derivative of the unknown growth function evaluated at time *d*.

### Hidden Markov Model (HMM) for growth stage classification

In order to automate the process of growth stage classification, we built a HMM that uses acceleration data obtained from MR as input. Considering **a** as the *n*-dimensional vector of estimated growth accelerations across *n* recorded days, we first compute **z** = sign(**a**), where sign(.) is a modified sign function which retrieves −1 for *a* < -*c*, +1 for *a* > *c* and 0 otherwise. Scalar *c* is defined as an acceleration cutoff, which is treated here as a free parameter. The objective of the HMM was to generate a sequence of states *K* = (*k*_1_, *k*_2_,…, *k*_n_) where each element *k*_i_ takes one of the following values: “lagging”, “exponential”, “deceleration” or “stationary”. The initial probabilities for these hidden states were set to 1, 0, 0 and 0, respectively, assuming that all growth curves start from a lagging stage. Now let **T** be a 4 × 4 matrix of transition probabilities between hidden states and **E** be a 4 × 3 matrix of emission probabilities that models the probability of each hidden state producing a *z* value of −1, 0 or +1. We adopted:

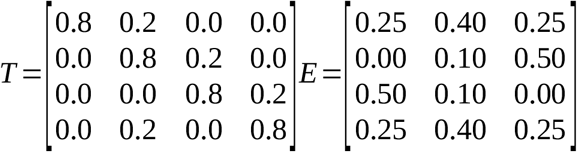

The selected values in **T** only permitted transitions lagging → exponential, exponential→ deceleration or deceleration → stationary. Values in **E** made *z* = 0 more likely to be produced by either the lagging or stationary stages, *z* = +1 more likely to be produced by the acceleration stage and *z* = −1 more likely to be produced by the deceleration stage. For the atypical transition deceleration → exponential, the described model would generate a short and intermediate stationary step between these two stages. In these cases, the spurious stationary step was replaced by an exponential classification after the HMM has been fitted to the data. The Viterbi algorithm implemented in the HMM v1.0 package (15) in *R* (9) was used to estimate the sequence *K*. After prediction of growth stages, stationary classifications were confronted against growth rates. If a given stationary stage presented a median growth rate greater than the maximum growth rate of the lagging phase, it was re-classified as a “linear” stage.

### Simulation study

To test the performance of MR in approximating growth curves and their rate of change and acceleration in scenarios where these curves have been observed only partially (i.e., real-time case report), we selected a widely used sigmoidal mathematical function, namely the Gompertz model, to generate 50,000 simulated growth curves. We used a parameterization of the Gompertz model that is dependent on four parameters:

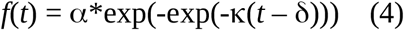

where *t* is a time point, α is the asymptote (i.e., number of cases at the stationary stage), exp is the exponential function, Κ is a growth coefficient and δ is the time at inflection of the exponential stage (i.e., time when the growth rate reaches its maximum value and acceleration transitions from positive to negative). All simulations were performed considering a 100-days period, with parameters sampled as follows: α ∼ Uniform(500, 10000), Κ ∼ Uniform(0.05, 0.95) and δ ∼ Uniform(5, 95). Completely stationary curves were discarded. The accuracy of growth rate and acceleration estimates produced by MR with smooth factor ranging from *s* = 3 to *s* = 10 were then evaluated by taking the coefficient of determination (*R*^2^) of the regression of true values onto estimates.

### Analysis of COVID-19 case reports

We analyzed case reports that have been updated daily by the European Center for Disease Prevention and Control (ECDC). The framework was applied to that data using smooth factors ranging from *s* = 3 to *s* = 10. The acceleration curves were clipped at observation *n* – *s* to avoid poor growth acceleration estimates at the end of the curve. Likewise, the last *s* days had their growth rates estimated by compounding rates from *n* – *s* to *n* using the acceleration estimated for day *n* – *s*. Finally, next-day predictions of COVID-19 prevalence were obtained by summing the last observed prevalence with its estimated growth rate. In order to measure the accuracy of these predictions, we performed a step-wise simulation by censoring observations ahead of each day, fitting MR to the remaining data and then comparing predicted and true next-day prevalence. Accuracy of predictions were again measured by linear regression.

### Analysis and visualization tools

All analyses presented in this paper were performed using *R* version 3.4.4 (9). To visualize the growth rate and acceleration of COVID-19 pandemic, we implemented a simple *Shiny* (8) dashboard application, which offers an intuitive web interface and allow us to be updated on new cases and the prevalence of COVID-19 worldwide. The application automatically loads the latest case reports from ECDC. Alternatively, users can upload their own data to visualize the growth rate and acceleration of COVID-19 of specific states, provinces, cities or aggregate data from arbitrary territory definitions. For the implementation we used the following packages: shiny v1.4.0 (16), shinydashboard v0.7.1 (17), shinydashboardPlus v0.7.0 (18), readxl v1.3.1 (19), shinyalert v1.0 (20), httr v1.4.1 (21) and plotly v4.9.2 (22), all available on CRAN (Comprehensive R Archive Network, https://cran.r-project.org/). The application can be downloaded from our GitHub repository at https://github.com/adamtaiti/SARS-CoV-2. A live instance of the app will be maintained until the end of the pandemic at https://www.theguarani.com.br/covid-19.

## Data Availability

The data in this study were obtained from the European Center for Disease Prevention and Control (ECDC) and are publicly available at https://opendata.ecdc.europa.eu/covid19/casedistribution/csv (accessed on April 2nd 2020). The source code for the R Shiny application used for data analysis is found in our GitHub repository: https://github.com/adamtaiti/SARS-CoV-2. A live instance of the app can be accessed at http://www.theguarani.com.br/.

## ACKNOWLEDGMENTS

We would like to express our highest gratitude to all health agents and individuals around the globe who were involved in reporting cases and making COVID-19 prevalence data available to the public. This study did not receive financial support and was conducted during voluntary social isolation.

## AUTHOR CONTRIBUTIONS

Y.T.U. conceived the study, performed simulations, coordinated the data analysis and wrote the manuscript. A.T.H.U. built R code for data analysis and programmed the Shiny App Dashboard. R.B.P.T., S.C.P., M.M. and J.F.G. revised growth curves for all countries/territories and pinpointed dates of measures taken by them to reduce human mobility. All authors revised and agreed with the contents of the manuscript.

## DATA AVAILABILITY

The data in this study were obtained from the European Center for Disease Prevention and Control (ECDC) and are publicly available at https://opendata.ecdc.europa.eu/covid19/casedistribution/csv (accessed on April 2^nd^ 2020). The source code for the *R Shiny* application used for data analysis is found in our GitHub repository: https://github.com/adamtaiti/SARS-CoV-2. A live instance of the app can be accessed at http://www.theguarani.com.br/covid-19.

